# The challenges of the coming mass vaccination and exit strategy in prevention and control of COVID-19, a modelling study

**DOI:** 10.1101/2020.12.18.20248478

**Authors:** Biao Tang, Peiyu Liu, Jie Yang, Jianhong Wu, Xiao Yanni, Sanyi Tang

## Abstract

With success in the development of COVID-19 vaccines, it is urgent and challenging to analyse how the coming large-scale vaccination in the population and the growing public desire of relaxation of non-pharmaceutical interventions (NPIs) interact to impact the prevention and control of the COVID-19 pandemic. Using mathematical models, we focus on two aspects: 1) how the vaccination program should be designed to balance the dynamic exit of NPIs; 2) how much the vaccination coverage is needed to avoid a second wave of the epidemics when the NPIs exit in stages. We address this issue globally, and take six countries--China, Brazil, Indonesia, Russia, UK, and US—in our case study. We showed that a dynamic vaccination program in three stages can be an effective approach to balance the dynamic exit of the NPIs in terms of mitigating the epidemics. The vaccination rates and the accumulative vaccination coverage in these countries are estimated by fitting the model to the real data. We observed that the required effective vaccination coverages are greatly different to balance the dynamic exit of NPIs in these countries, providing a quantitative criterion for the requirement of an integrative package of NPIs. We predicted the epidemics under different vaccination rates for these countries, and showed that the vaccination can significantly decrease the peak value of a future wave. Furthermore, we found that a lower vaccination coverage can result in a subsequent wave once the NPIs exit. Therefore, there is a critical (minimum) vaccination coverage, depending on effectiveness of NPIs to avoid a subsequent wave. We estimated the critical vaccination coverages for China, Brazil, and Indonesia under different scenarios. In conclusion, we quantitatively showed that the dynamic vaccination program can be the effective approach to supplement or even eventually replace NPIs in mitigating the epidemics and avoiding future waves, and we suggest that country level-based exit strategies of the NPIs should be considered, according to the possible quarantine rate and testing ability, and the accessibility, affordability and efficiency of the vaccines.

## Introduction

Since January 23, 2020, in combating with the COVID-19 epidemics, the Chinese government adopted a series of non-pharmaceutical interventions (NPIs), including lockdown of the city, close contact tracing and quarantine, isolation, enhanced testing^1^. With this package of NPIs, mainland China has reporting no local COVID-19 outbreaks since March 19th, 2020, supporting the high effectiveness of these NPIs^2-4^. Many other countries carried similar strategies in mitigating the COVID-19 pandemic, with varying outcomes. In parallel, many modelling studies quantitatively demonstrated that NPIs implemented were and are effective in mitigating the COVID-19 epidemics^5-9^. However, many countries failed to prevent subsequent waves even with higher peaks than that of the first wave after the reopening^10^.

COVID-19 vaccination has been believed to be the potentially most feasible method to eventually put the pandemic under control. Reported from the WHO, there are over 169 COVID-19 vaccine candidates under development, with 26 of these in the human trial phase as of October 2, 2020^11^. Several modelling studies^12-14^ have tried to investigate the effectiveness of COVID-19 vaccination in mitigating the epidemics, several of which focus on the optimal control of the vaccination in mitigating the epidemics^15-19^. However, the vaccine supply is limited and the first 10 million to 15 million doses may be enough to cover around 3% to 5% of the U.S. population, from the estimates of the government’s vaccine project^20^. The COVID-19 vaccine would be much less in the developing countries. With the limited COVID-19 vaccine supply, it remains a challenge how to design an optimal COVID-19 vaccination regime and estimate how many doses are needed to avoid a subsequent wave with the relaxed NPIs after re-opening. Taking six countries (China, Brazil, Indonesia, Russia, UK, and US) with different types of epidemic curves as a case study, we aim to address these critical issues using a transmission dynamics model.

The main purpose of this study is to use mathematical models to quantitatively discuss how a dynamical vaccination program should be designed and how many doses are needed to replace the existing NPIs in terms of mitigating the epidemics during an outbreak, and also to avoid subsequent waves after reopening with relaxed NPIs. The rest is organized as follows. In the coming section, we first propose the compartment models of COVID-19 transmission dynamics with and without vaccination. Also, we provide the preliminary methodologies in this section. In section 3, we calibrate the model without vaccination by fitting the COVID-19 epidemic data, and estimate the unknown parameters for the six countries. Further, by re-fitting the model with vaccination to the same data, we evaluate if the pre-designed vaccination program can balance the exits of the NPIs in different countries, and what the vaccination speed should be. In section 4, we investigate how the vaccination can mitigate the epidemic and how much vaccination coverage is needed to avoid a subsequent wave. Finally, we make the conclusion and mark the important points in the vaccination progress.

## Methods

### Data

We obtained the data of daily COVID-19 confirmed cases and daily death cases linked to COVID-19 in China, Brazil, Indonesia, Russia, UK, and US from the website https://github.com/CSSEGISandData/COVID-19, as shown in Fig. 1. Note that, except China, we included a time series with a same length in the rest five countries (i.e. 210 days). Based on the data information included, the six countries can be divided into two subgroups. The one group includes China, Brazil, and Indonesia, who experienced one epidemic wave till Oct. 13, 2020. In details, China has successfully controlled the epidemic with the first wave being finished already, the epidemics in Brazil or Indonesia are in the decreasing phase. The other group includes Russia, UK, and US, who experienced two epidemics waves till Oct. 13, 2020. The epidemics of UK and Russia are in the increasing phase while the epidemic of US belongs to the decreasing phase till the last data point of Fig. 1. It should be mentioned that US is actually experiencing the third wave of the COVID-19 epidemic till December 04, 2020, which can be a validation for our prediction results. The data were released and analysed anonymously.

**Figure 1.**
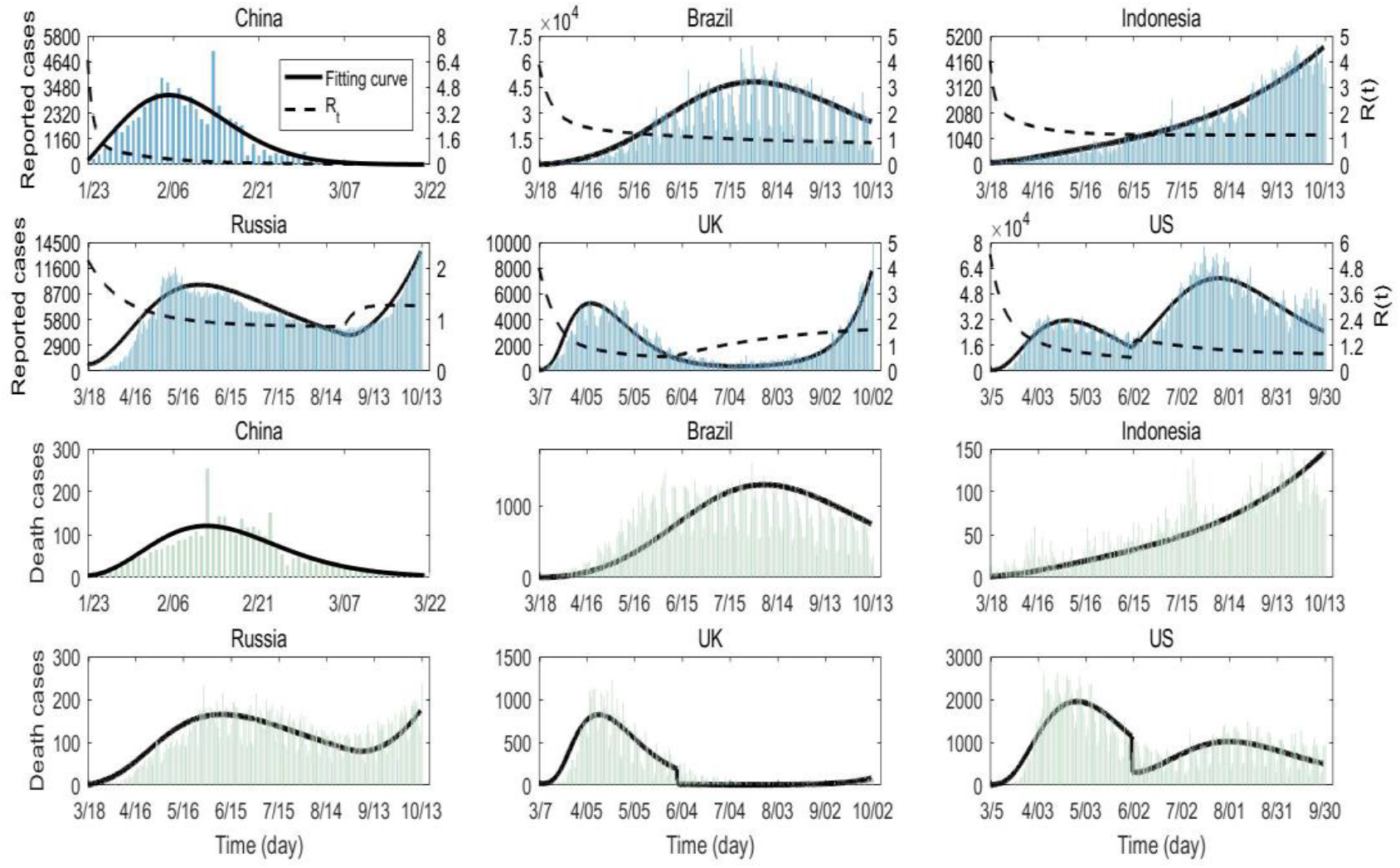
COVID-19 epidemic data and best fitting results in China, Brazil, Indonesia, Russia, UK, and US. The upper two panels are the daily reported cases in the six countries, while the lower two panes are the daily death cases in the six countries. The solid black curves are fitted solutions of model (1) while the dash black curves are the corresponding effective reproduction number *R*_*t*_.

### The models

We firstly develop a modelling framework of COVID-19 transmission dynamics for the six countries based on the previous modelling studies^21-23^. The population is divided into susceptible (*S*), exposed (*E*), asymptomatic infectious (*A*), infectious with symptoms (*I*), and recovered (*R*) compartments according to the epidemiological status of individuals, and further into diagnosed and hospitalized (*H*), quarantined susceptible (*S*_*q*_), and isolated exposed (*E*_*q*_) compartments based on control interventions. We also account for contact tracing, where a proportion, *q*, of individuals exposed to the virus are quarantined. The quarantined individuals can either move to the compartment *E*_*q*_ or *S*_*q*_, depending on whether they are effectively infected or not, while the other proportion, 1 – *q*, consists of individuals exposed to the virus who are missed from contact tracing and, therefore, move to the exposed compartment *E* once effectively infected, or stay in the compartment *S* otherwise (6). The dynamics model is given by:

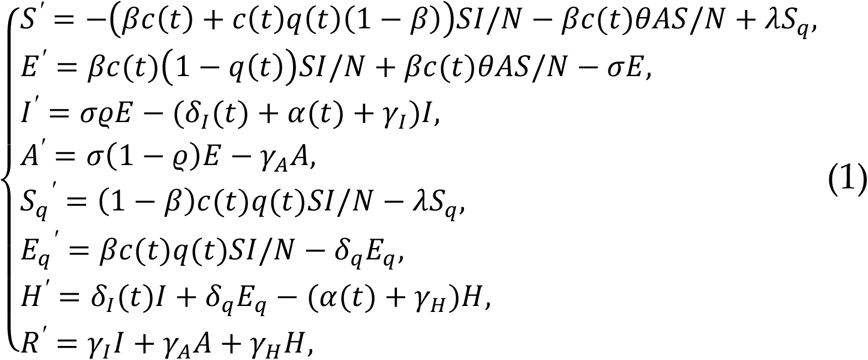

where, *N*=*S*+*E*+*I*+*A*+*Sq*+*Eq*+*H*+*R* is the total population.

Considering the continuously enhanced NPIs, we set the quarantine rate, the diagnose rate, and the contact rate as a function of time *t*. In details, the quarantine rate and the diagnose rate is increasing gradually with the following forms^22,23^

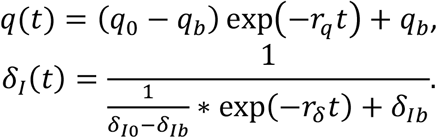

For the countries with one epidemical peak, we assume that the contact rate is decreasing overtime, which is given by

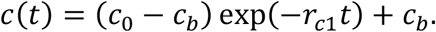

For the countries with two epidemic waves, we assume that the contact rate should first decrease because of the implementation of NPIs and increase again since the reopening. Therefore, the contact rate in Russia, UK, and US, is of the following form

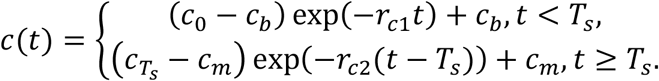

where 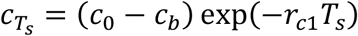 is the contact rate at time *T*_*s*_, and *c*_*m*_is the potential maximum contact rate after reopening with *c*_*m*_ < *c*_0_. *T*_*s*_ is the initial time of the reopening. Note that, the UK government announced to reopen the market and the schools gradually starting at June 1, 2020, thus we set *T*_*s*_as June 1, 2020 for UK. However, there is no definite time point to start the reopening in Russia and US. There are two reasons, one is that the NPIs are enhanced or weakened in turn, the other one is that the initial time of the reopening in different states of the country are greatly different. Therefore, based on the real data, we assume that the initial time of the reopening is the day with smallest number of reported cases between the two waves. Consequently, *T*_*s*_is chosen as August 25, 2020 and May 31, 2020 in Russia and US, respectively. It should be mentioned that for countries with more than two epidemic waves (such as the epidemic in United States till December 04, 2020), we can use the same methods to define the above time-dependent rates.

For the purpose to better fit the model to the data of death cases linked to COVID-19, we set the disease-induced death rate as a piecewise function of time *t* in US and UK. For simplicity, we assume that the death rate changed after the reopening. Thus, there is

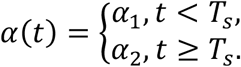

The death rates in the rest four countries are assumed to be constant during the time interval included. All the other parameters are positive constants. The definitions of all the variables and parameters are listed in Table 1.

**Table 1:**
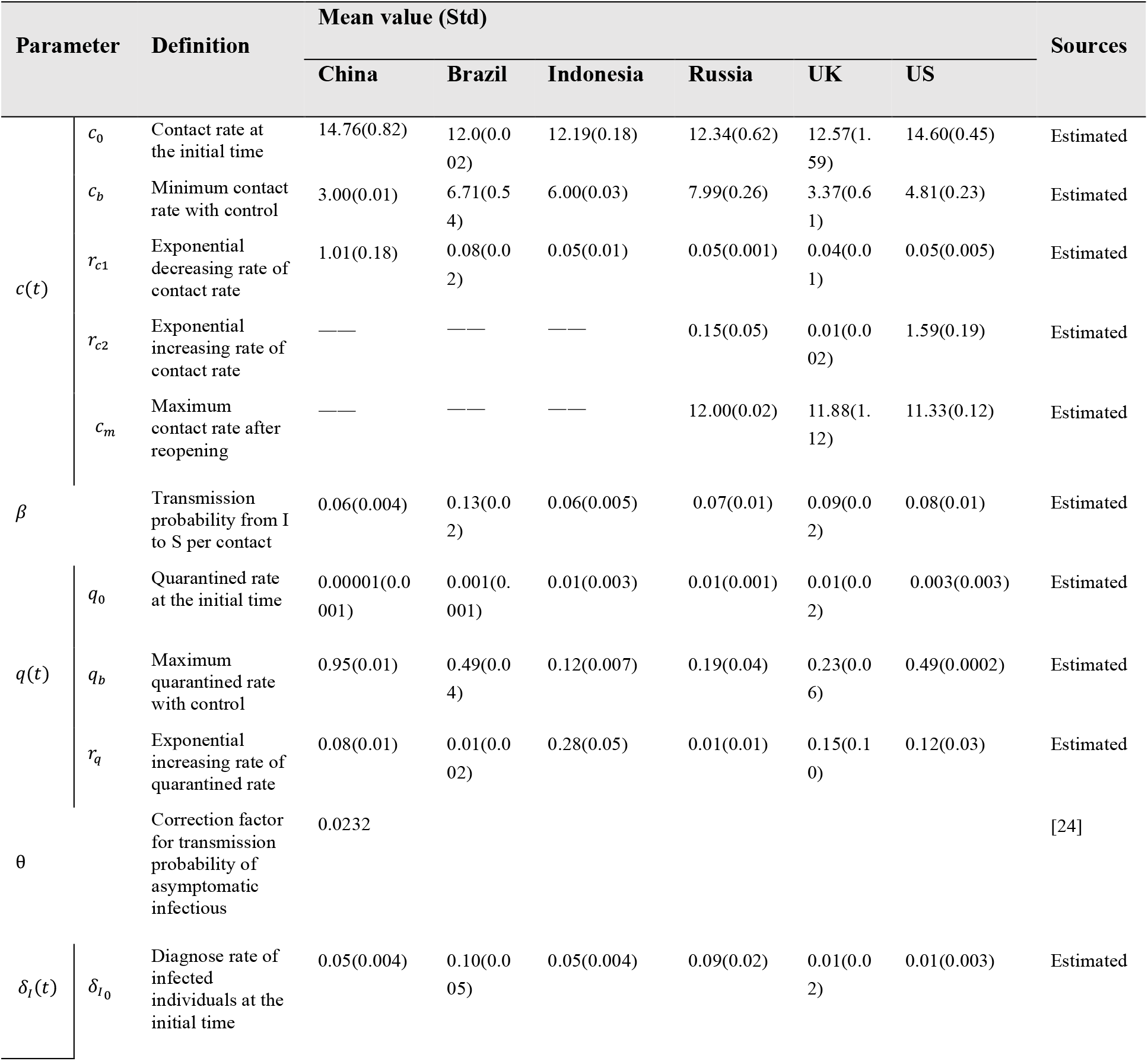
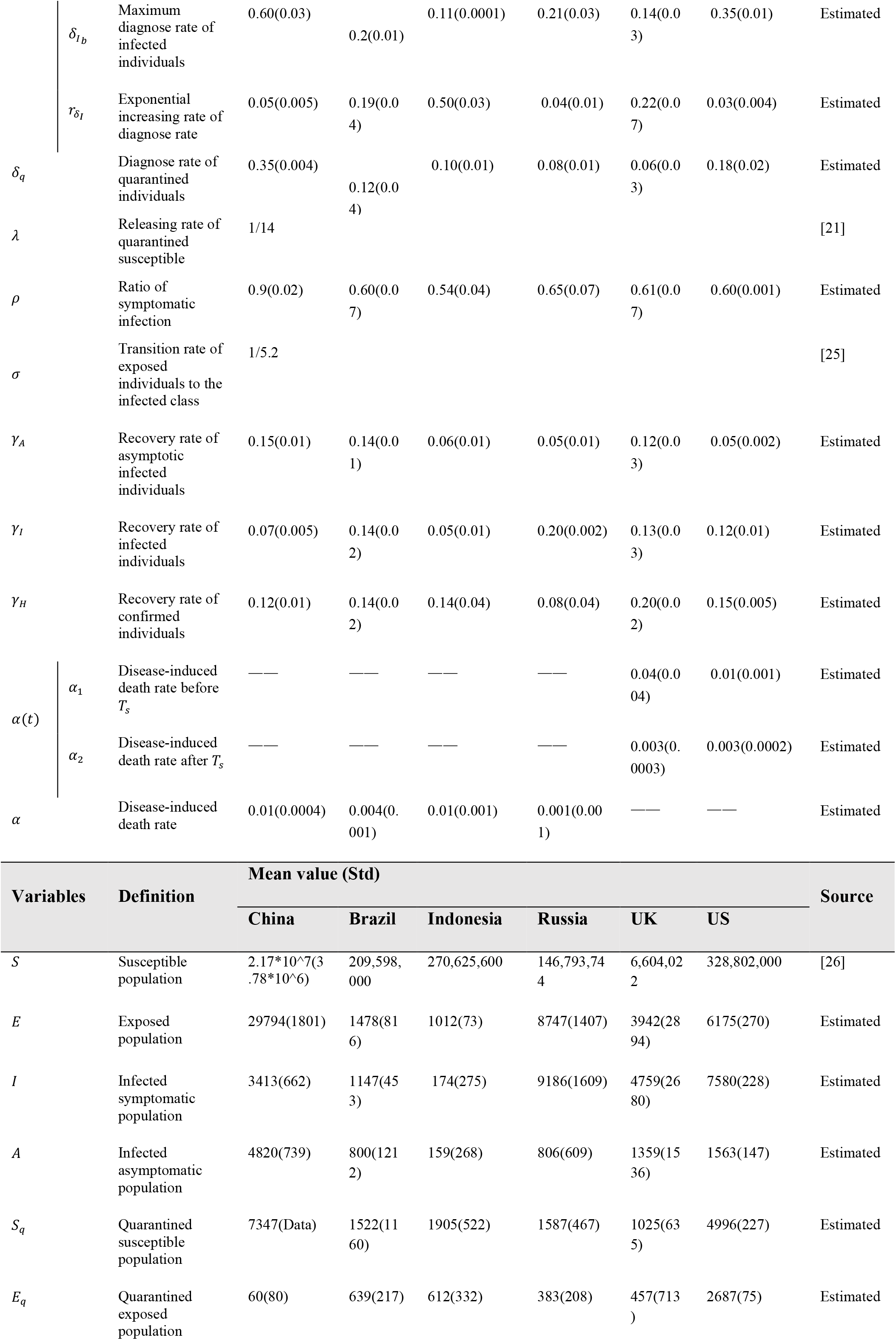
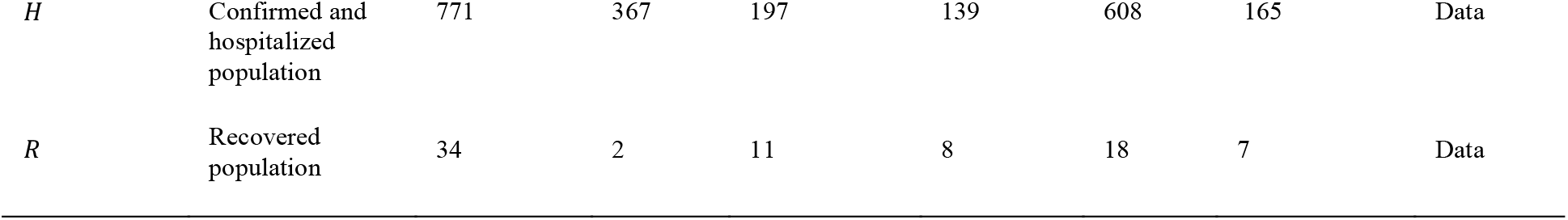
Parameter estimates of model (1) in the six countries.

In addition, if a COVID-19 vaccine is available, a vaccination schedule in multiple stages is considered, and each stage has a different vaccination speed (i.e. vaccination rate). Incorporating this kind of vaccination regime, then model (1) becomes:

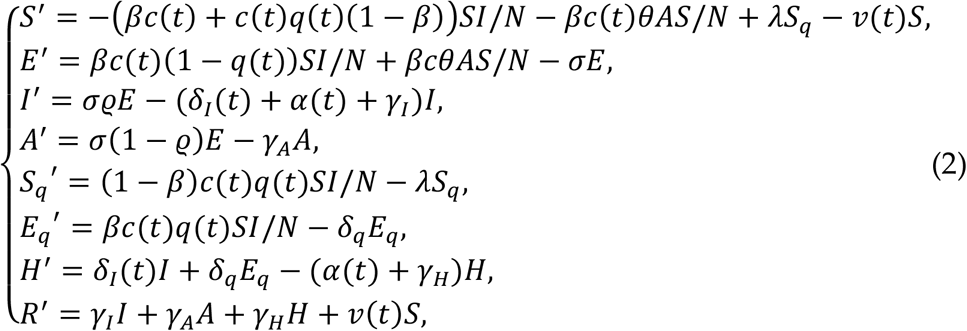

where *v*(*t*) is a piecewise continuous function of time *t*, which denotes the time-dependent (dynamic) vaccination rate of COVID-19.

As we mentioned in the introduction, we mainly consider two vaccination regimes in the population. Firstly, during the outbreak, we vaccinate against COVID-19 to balance the exit of NPIs also starting at time *T*_*v*_in three stages, and each stage lasts for *τ* days with a constant vaccination rate *v*_*i*_, *i =* 1,2,3. Therefore, the vaccination rate *v*(*t*) is of the following form:

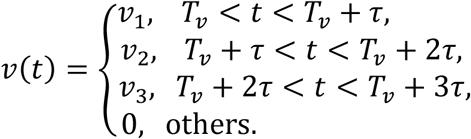

The effective accumulative vaccination coverage can be defined as:

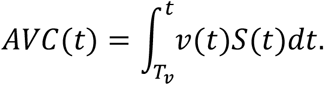

Particularly, we denote 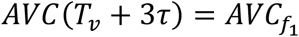

To conduct the sensitivity analysis of the starting time of the vaccination, we choose three time points of *T*_*v*_, basically which lie in the increasing phase, the peak time, and the decreasing phase of the epidemics, respectively. The detailed starting time in the six countries are given in Table 2. Furthermore, as the epidemical period in China is much shorter than those in the other five countries, we assume that the vaccination last for 7 days in each stage in China, i.e. τ=7, while it lasts for 15 days in each stage in the rest five countries with τ=15.

**Table 2:** Estimated vaccination rate and coverage needed to balance the dynamic exit of NPIs.

| Country | $T_v$ | $a_1$ | ES1 | | | ES2 | | | | |
| --- | --- | --- | --- | --- | --- | --- | --- | --- | --- | --- |
| | | | $v_1$ | $v_2$ | $v_3$ | $AVC_{f_1}$ | $v_1$ | $v_2$ | $v_3$ | $AVC_{f_1}$ |
| China | 8 | 1.6989 | 0.155 | 0.013 | 0.07 | 0.7878 | 0.07 | 0.42 | 0 | 0.9592 |
|  | 13 | 1.7007 | 0.155 | 0.027 | 0.035 | 0.7735 | 0.105 | 0.43 | 0 | 0.9675 |
|  | 25 | 1.7007 | 0.135 | 0.0103 | 0.001 | 0.6420 | 0.12 | 0.40 | 0 | 0.9660 |
| Brazil | 80 | 1.2132 | 0.025 | 0.006 | 0.0074 | 0.4271 | 0.035 | 0.019 | 0.011 | 0.6068 |
|  | 130 | 1.2137 | 0.0305 | 0.0003 | 0.007 | 0.4096 | 0.04 | 0.019 | 0.0105 | 0.6207 |
|  | 165 | 1.2137 | 0.0289 | 0.0018 | 0.0091 | 0.4181 | 0.042 | 0.015 | 0.02 | 0.6498 |
| Indonesia | 80 | 1.2560 | 0.0361 | 0.0051 | 0.0052 | 0.4994 | 0.029 | 0.02 | 0.0084 | 0.565 |
|  | 130 | 1.2658 | 0.0366 | 0.002 | 0.0052 | 0.4922 | 0.0285 | 0.0174 | 0.0125 | 0.5751 |
|  | 165 | 1.2666 | 0.0343 | 0.004 | 0.006 | 0.4861 | 0.0281 | 0.0177 | 0.0128 | 0.5855 |
| Russia | 32 | 1.1158 | 0.017 | 0.008 | 0.003 | 0.3458 | 0.016 | 0.0165 | 0.003 | 0.4176 |
|  | 68 | 1.1494 | 0.018 | 0.008 | 0.003 | 0.344 | 0.016 | 0.019 | 0.002 | 0.4283 |
|  | 100 | 1.1549 | 0.018 | 0.007 | 0.003 | 0.3401 | 0.016 | 0.02 | 0.002 | 0.4214 |
| **US** | 43 | 1.3253 | 0.048 | 0.005 | 0.05 | 0.7593 | 0.05 | 0.045 | 0.05 | 0.8746 |
|  | 142 | 1.0881 | 0.0113 | 0.0024 | 0.0055 | 0.2319 | 0.0327 | 0.021 | 0.001 | 0.5432 |
|  | 165 | 1.0881 | 0.0113 | 0.0021 | 0.0055 | 0.2336 | 0.031 | 0.021 | 0.001 | 0.5287 |
| **UK** | 15 | 1.1238 | 0.046 | 0.0026 | 0.02 | 0.6312 | 0.042 | 0.029 | 0.0033 | 0.6655 |
|  | 34 | 1.2731 | 0.049 | 0.014 | 0.008 | 0.6519 | 0.049 | 0.019 | 0.012 | 0.6943 |
|  | 165 | 1.1282 | 0.01 | 0.006 | 0 | 0.208 | 0.0168 | 0.016 | 0 | 0.3886 |

Note that, ‘AVR’ means the effective accumulative vaccination coverage. ES1 and ES2 are the two exit strategies defined above.

Secondly, we assume to vaccinate against COVID-19 starting at the last date of the real data in each country, denoted*T*_*e*_, and the vaccination lasts a period of 21 days with a constant rate. Then, there is

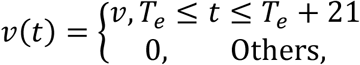

where *v >* 0. In this case, the effective accumulative vaccination coverage is given by

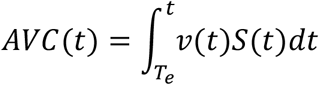

with 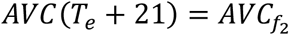.

### Exit strategy

To analyse how to use a COVID-19 vaccine in the population to balance the dynamic exit of the NPIs, we consider two types of exit strategy. Aiming at alleviating the urgent needs to resume the normal-life, we assume that the population can resume the daily-life activity (in terms of the contact rate) in three stages in consistent with the vaccination regimes. Consequently, the contact rate has the following form:

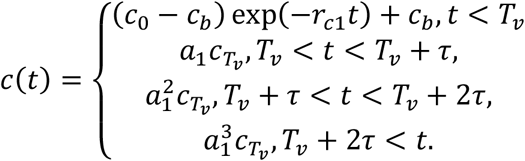

where 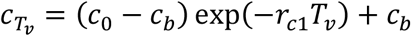 is the contact rate at time *T*_*v*_. Here, we assume that the contact rate can return to the initial value in the three stages at the same rate, i.e. 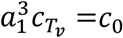 Note that, when the population resume the normal-life activity, we can still remain the normalized control and prevention by keeping the enhanced quarantine rate and diagnose rate. We call this exit strategy that the contact rate returns to the initial value but the quarantine rate and the diagnosed rate remain the estimated values as “ES1”.

On the other hand, at the same time as the population resume their normal-life, we also release the control intervention of close contact tracing and quarantine. That is, we assume that the quarantine rate becomes zero since the beginning of vaccination. Thus, the quarantine rate is of the following form

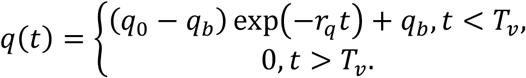

The exit strategy that the contact rate return to the initial value and the quarantine rate becomes zero is called as “ES2”.

When we project the epidemics in the six countries by using a vaccine to mitigate and avoid a second wave of COVID-19 epidemics when the NPIs exit. Differ to the above case, we assume that the population return to a normal life-activity starting at the end point of the time series of real data, denoted by *T*_*e*_. That is, the contact rate return to *c*_0_directly. Thus, there are

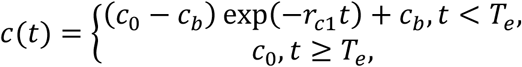

for the countries with one epidemic peak, and

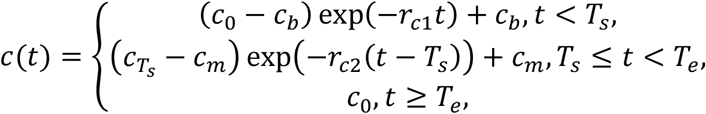

for the countries with two epidemic peaks.

Similarly, we also consider two scenarios in terms of the quarantine rate. That is, ES1: the quarantine *q*(*t*) remains the estimated values; ES2: the quarantine rate *q*(*t*) is set to be 0 after time *T*_*e*_ with

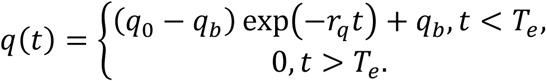

## Main results

### Model calibration

To estimate the parameters of model (1), we initially informed parts of the parameters in model (1) with fixed values from existing studies or database, as listed in Table 1. We used a bootstrap method to generate 500 time series of both the reported cases and death case from a Poisson distribution with mean given by the real data. Then, we fitted the model to each dataset of the time series of the reported cases and the death cases simultaneously in each country. Based on the 500 times fitting results, we obtained the mean and their standard deviations of all unknown parameters, which are listed in Table 1. Using the estimated mean values of all the parameters, we plotted the best fitting results of model (1) for the six countries, i.e. the solid curves marked as black in Fig. 1.

### The vaccination program to balance the dynamic exit of NPIs

In this section, we estimate the vaccination rate *v*_*i*_, *i =* 1,2,3 to balance the dynamic exit of the NPIs defined in the method section. All the parameters, except for the quarantine and the contact rate, are fixed as the same as those of the calibration results of model (1). Based on the estimated results by fitting model (1) to the data, we can also calculate the values of *a*_1_for six countries, which are listed in Table 2.

Incorporating the assumptions in the method section and using the least square method, we fitted model (2) (i.e. the model with COVID-19 vaccination) to the time series of both the reported cases and death case for six countries. Note that as for the countries with two peaks, we only fitted one of the waves. In details, we only fitted the data before *T*_*s*_ when *T*_*s*_ < *T*_*v*_, and we fitted the data after *T*_*s*_ when *T*_*s*_ *> T*_*v*_. The best fitting results for China, Brazil, and Indonesia are shown in Fig. 2 while the best fitting results for Russia, US, and UK are shown in Fig 3. Table 2 also provided the estimated values of the vaccination rates in the three stages and the corresponding effective accumulative vaccination coverage 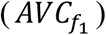 in different scenarios for six countries.

**Fig. 2.**
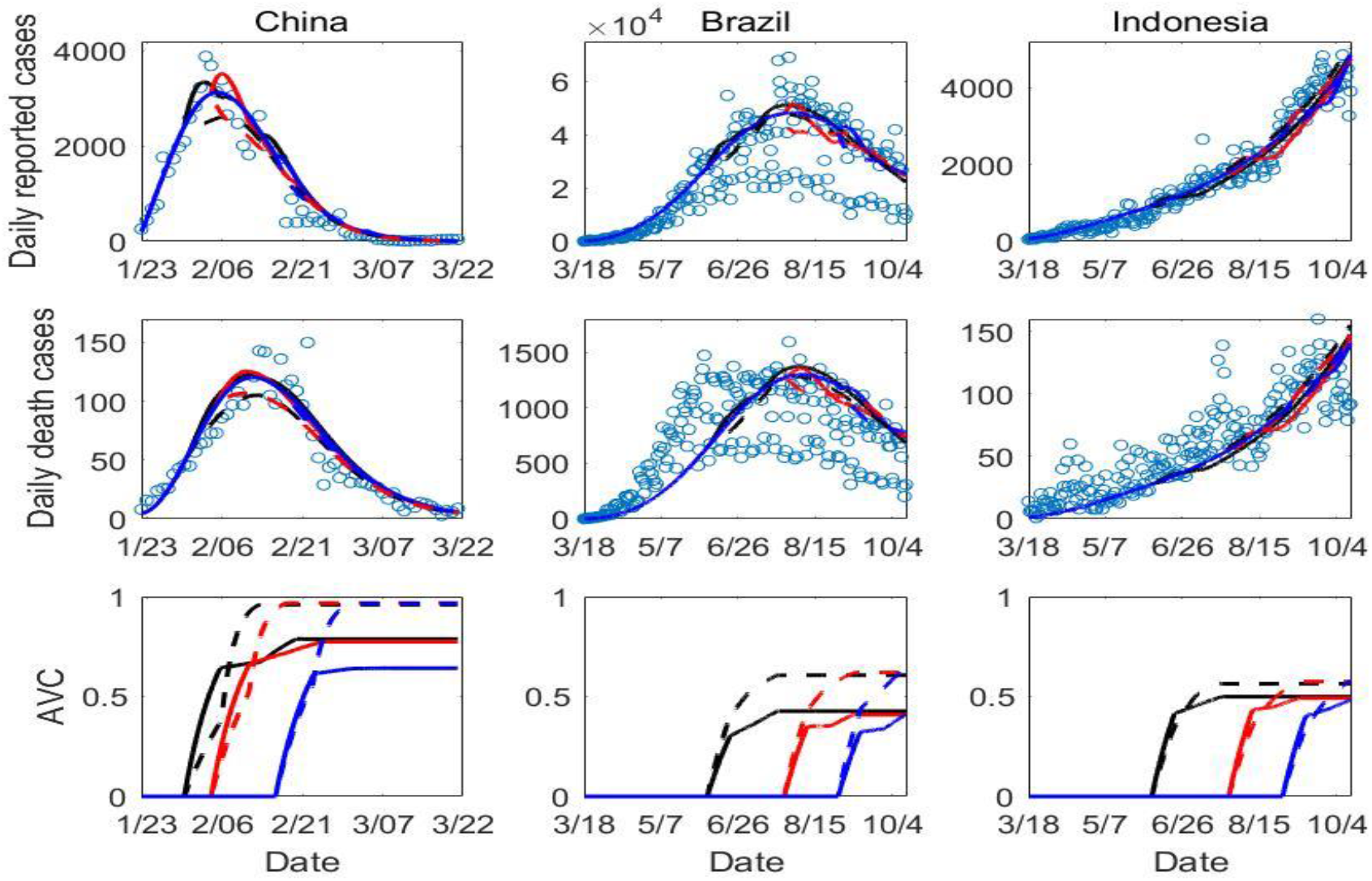
The balance results for the countries with one epidemic wave. The different colours denote the different starting time of the vaccination (*T*_*v*_) as listed in Table 2. The dash lines are the results under the exit strategy of ES2 while the solid lines are the results under the exit strategy of ES1. The cycles are the real data.

**Fig. 3.**
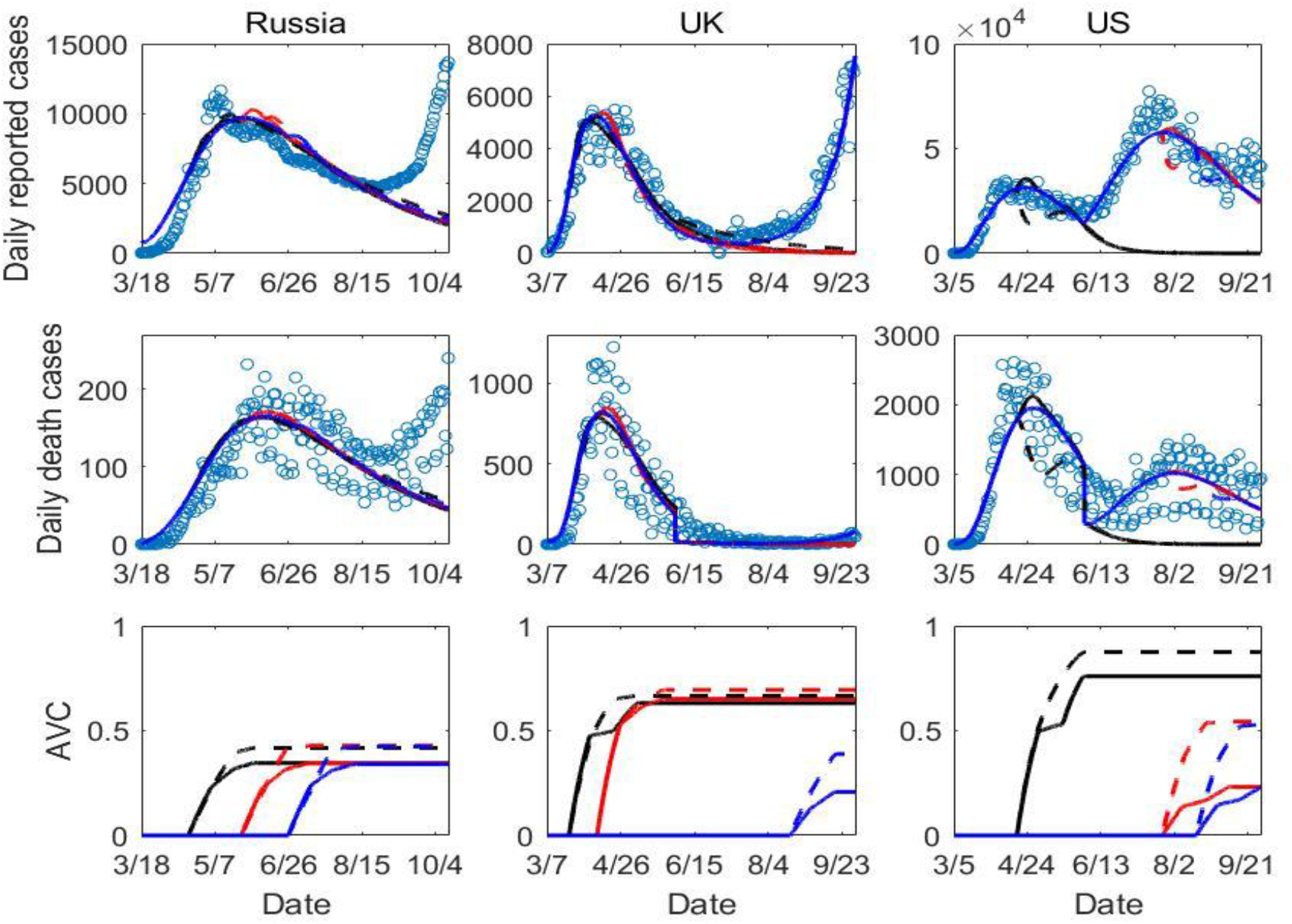
The balance results for the countries with two epidemic waves. The different colours denote the different starting time of the vaccination (*T*_*v*_) as listed in Table 2. The solid curves are the results under the exit strategy of ES2 while the dash curves are the results under the exit strategy of ES1. The cycles are the real data.

It follows from Figs.2-3 that the model with vaccination (i.e. model (2)) can fit the real data very well, which indicates that the designed dynamic vaccination program can be an effective method to replace the role of NPIs in terms of mitigating the COVID-19 epidemics. From Table 2, we can see that the values of the vaccination rates in the three stages are of a decreasing trend. Therefore, a fast vaccination in the early stage is required to control the outbreaks. Furthermore, with three different starting times of the vaccination (different colours in Figs.2-3), the best fitting curves can always have a similar shape to fitting results of the model without vaccination (i.e. the solid curves in Fig.1) in the same scale. Thus, whenever the vaccine is available during an outbreak, vaccination can be carried out to replace the NPIs aiming at controlling the outbreak.

Comparing the dash lines or solid lines in different colours of Fig. 2, it’s interesting to note that the accumulative vaccination coverage tends to be similar for the countries with a single epidemic wave (i.e. China, Brazil, and Indonesia) no matter when we start to vaccinate the population. In contrast, as for UK and US with two epidemic waves, it follows from Fig. 3 and Table 2 that the accumulative vaccination coverage for *T*_*v*_ *> T*_*s*_ is much lower than those for *T*_*v*_ < *T*_*s*_. A further comparison of the dash line and the corresponding solid line in the last row of Fig. 2, we observed that the accumulative vaccination coverage 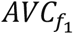 in the exit strategy of ES1 is lower than those in the exit strategy of ES2. This means that relatively low level of vaccination coverage can also successfully control an outbreak by combining with the NPIs.

Further, high vaccination coverage to balance the exit of the existing NPIs indicates the relatively high strengthen of the control interventions in the corresponding country. Based on this fact, we can conclude that: 1). the vaccination coverage in China ranked as the first among these six countries, quantitatively supporting that the combined control measures implemented in China were the strictest; 2). comparing the accumulative vaccination coverages in the exit strategy of ES1 and ES2, there is only a slight difference between the two coverages (less than 0.1) for Indonesia, Russia, and UK, which implies that the strength of the interventions is relative weak in these three countries; 3). the control strengthen becomes weaker and weaker during the period of the second wave compared with those with the first wave in US and UK, as a lower vaccination coverage is estimated when *T*_*v*_ *> T*_*s*_ as shown in Fig. 3. In other words, if one cannot successfully control an emerging infectious disease in a short period, it’s hard to avoid a more serious second wave with a larger peak as it is impossible to persistently implement the strengthened control interventions.

Next, taking China as an example, we conduct the sensitivity analysis to show how the starting timings of the vaccination and the vaccination speed in different stages affect the COVID-19 epidemics. In Fig. 4 and Fig. 5, we plotted the daily reported cases of model (2) by decreasing the values of vaccination rates (*v*_*i*_, *i =* 1,2,3) in the exit strategy of ES1 and ES2, respectively. From Fig. 4(b), we find that vaccination against COVID-19 initiating from the increasing phase of the epidemic has a significant impact on the epidemics, that is, a great vaccination rate will result in a low peak. However, if the vaccination is initiated at the decreasing phase, we observe that vaccination has a limited influence on the epidemics, i.e. the difference of the epidemic curves with different vaccination rate are tiny even when the AVC increased around 3 folds, shown in Fig. 4(g). We can obtain the similar results by comparing the first row and the third row of Fig. 5. Differ to the results in Fig. 4, we observed in Fig. 5 that if the vaccination rate is small (or the vaccination coverage is low), a second wave of the epidemics (even with a larger peak) may occur with the exit strategy of ES2. This indicates the necessity of persistently keeping the normalized interventions if we have a limited access to vaccine or the low efficacy of the vaccine.

**Fig. 4.**
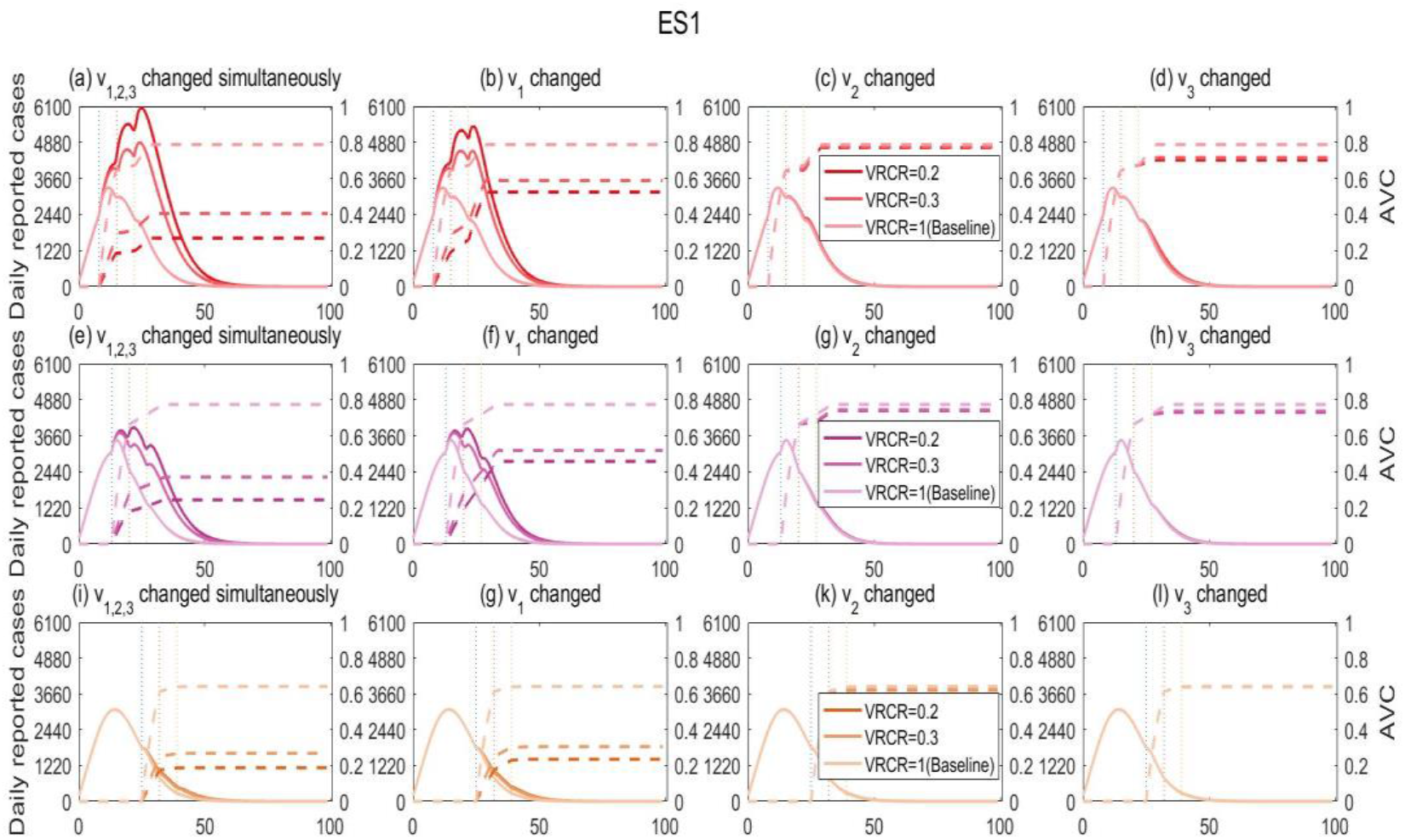
Sensitivity analysis. Solutions of model (2) for China with different vaccination rates under the exit strategy of ES1 defined in the method section. (a)-(d) *T*_*v*_ *=* 8; (e)-(h) *T*_*v*_ *=* 13 (i)-(l) *T*_*v*_ *=* 25.Here, ‘AVC’ denotes the effective accumulative vaccination coverage defined in the method section, ‘VRCR’ is the changing ratio of vaccination rate. ‘VRCR=1’ means that the vaccination rate (*v*_*i*_, *i =* 1,2,3)are chose as the estimated value while ‘VRCR=1’ means that the vaccination rates are the half of the estimated value. The dash lines are the AVC corresponding to the vaccination rate with the same colour.

**Fig. 5.**
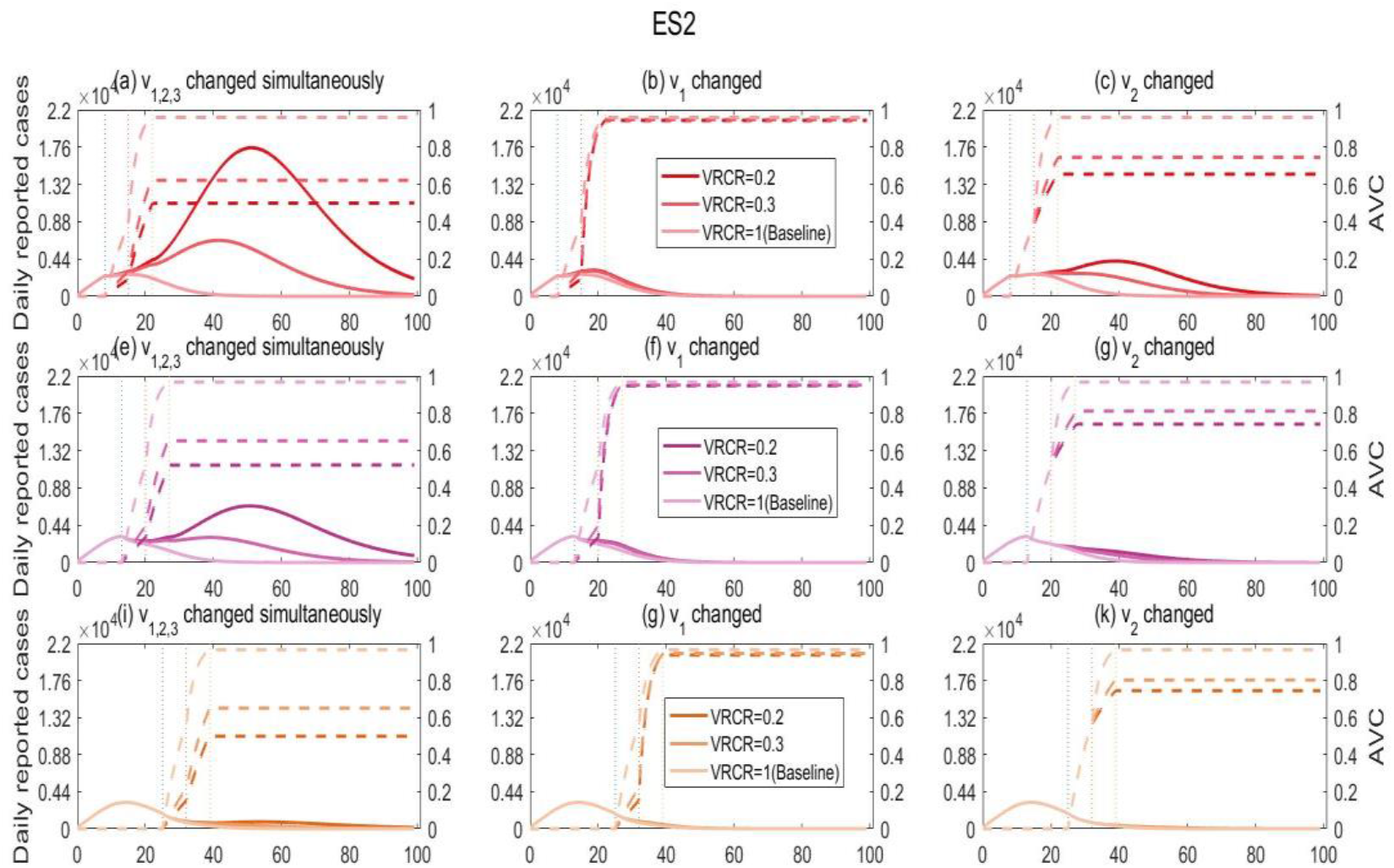
Sensitivity analysis. Solutions of model (2) for China with different vaccination rate under the exit strategy of ES2 defined in the method section. (a)-(c) *T*_*v*_ *=* 8; (e)-(g) *T*_*v*_ *=* 13(i)-(k) *T*_*v*_ *=* 25.The other settings are same to those of Fig. 4.

### Projection of vaccination to mitigate and avoid a second wave of COVID-19 epidemics

Based on the assumptions in the method section, we predicted the epidemics in the six countries with different vaccination rates. In Fig.6 and Fig.7, we plotted the daily confirmed cases under the exit strategy of ES1 and ES2, respectively. In order to make a comparison, we also predicted the COVID-19 epidemics with different vaccination rates by keeping all the control interventions (i.e. no exit), as shown in Fig.8. Table 3 provided the effective accumulative vaccination coverage in different scenarios of Figs. 6-8 and the corresponding peak values of the daily reported cases since the last date of the real data.

**Table 3:**
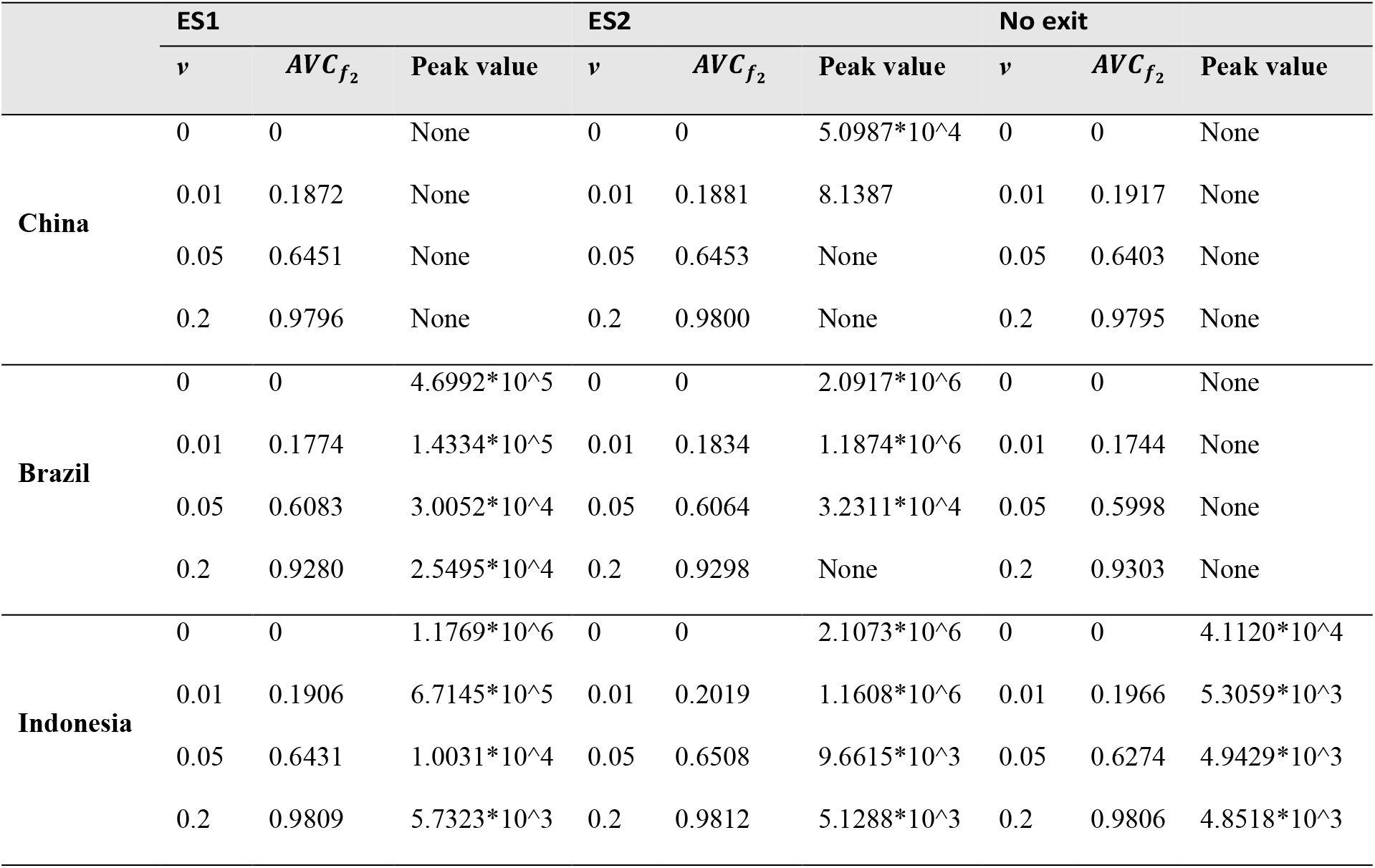
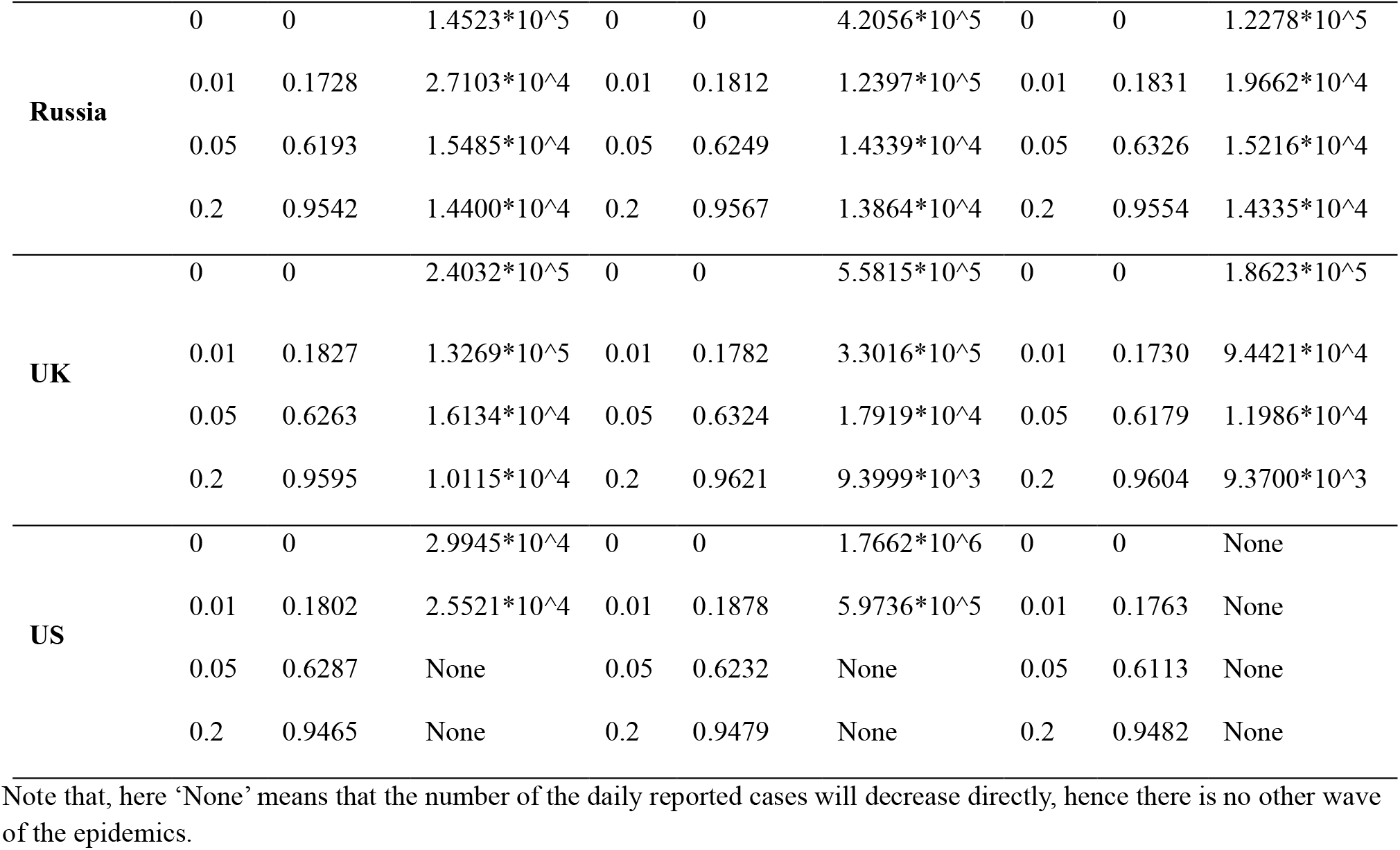
Prediction of the peak values of the daily reported cases.

**Fig. 6.**
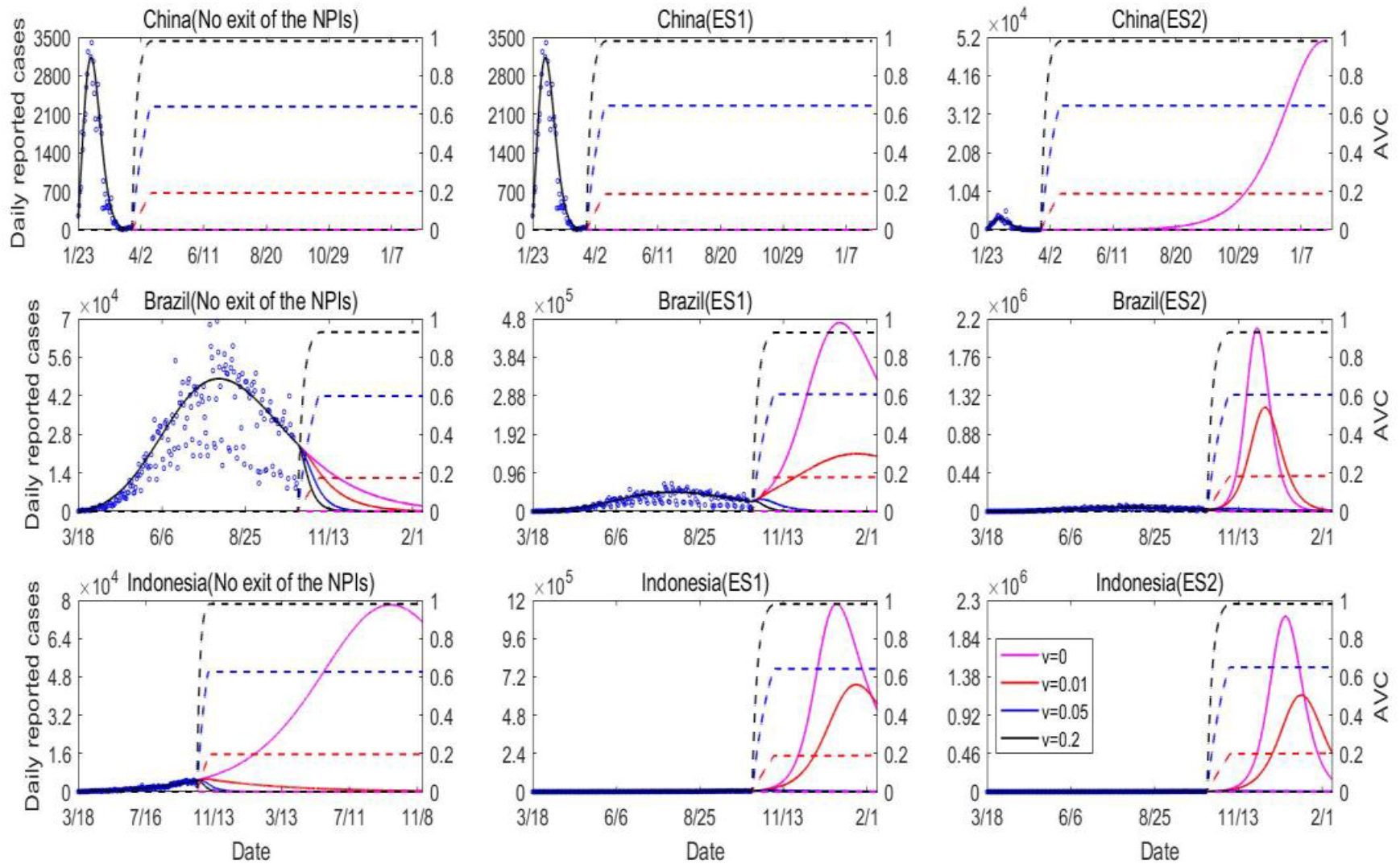
Predictions of the COVID-19 epidemics in three countries with one epidemic wave by solving model (2) with different vaccination rate and in different exit strategies. The dash lines are the corresponding AVC with the same colour of the vaccination rates.

**Fig. 7.**
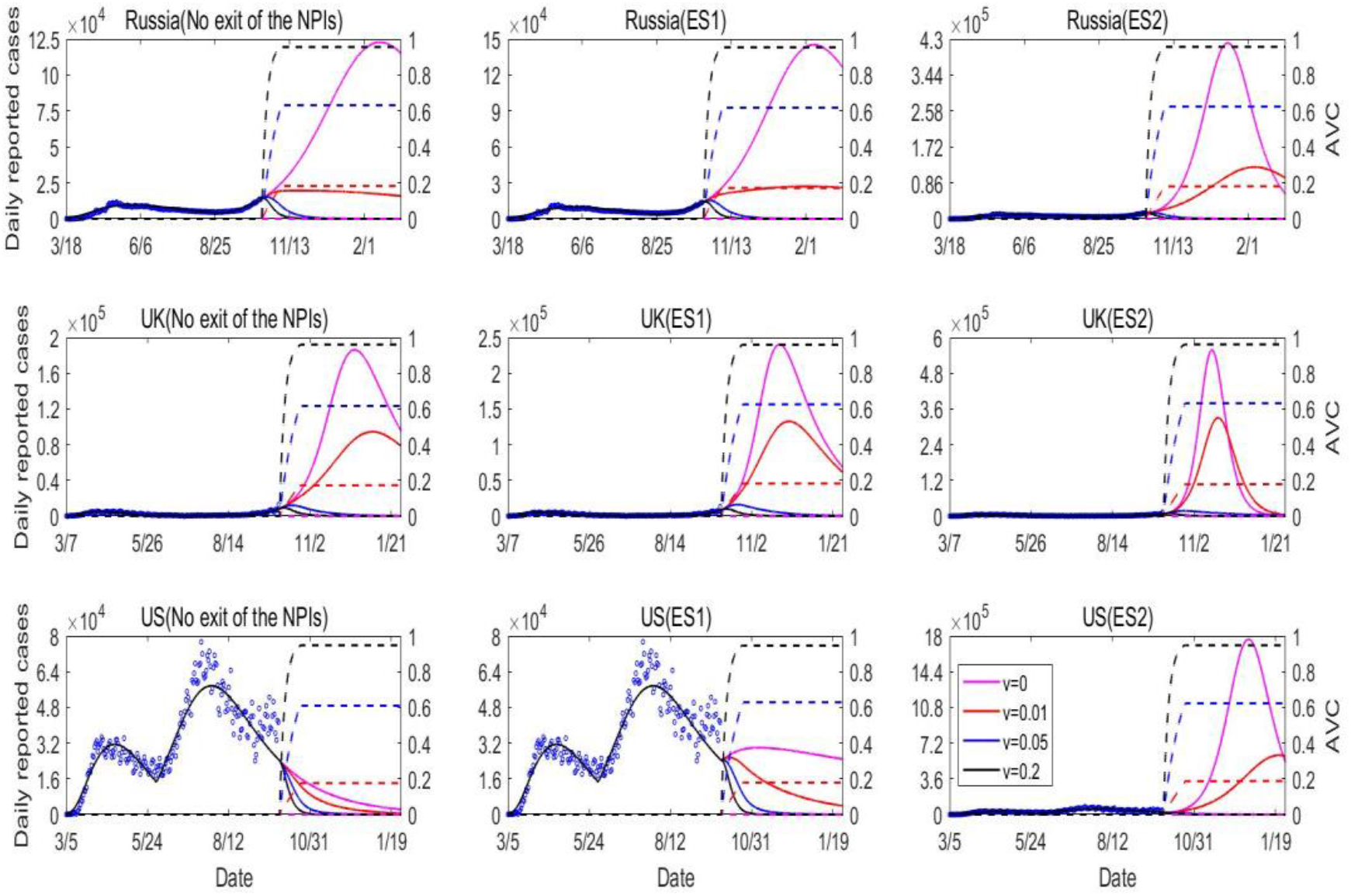
Predictions of the COVID-19 epidemics in the three countries with two epidemic waves by solving model (2) with different vaccination rate and in different exit strategies. The dash lines are the corresponding AVC with the same colour of the vaccination rates.

**Fig. 8.**
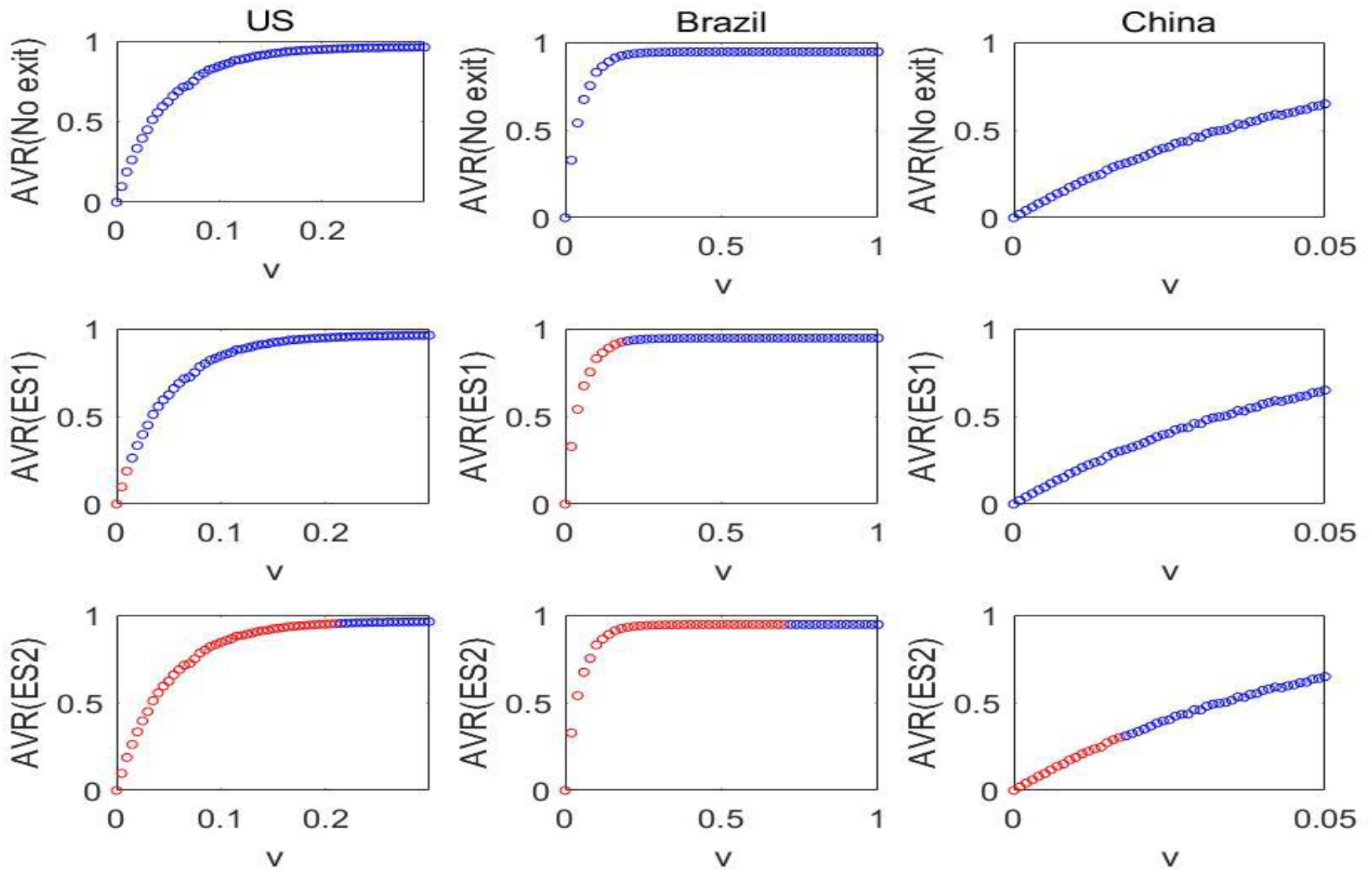
Relation of the vaccination rate and the accumulative vaccination coverage. Here, we take the newly infection by solving model (2) as the index to judge if there is a second wave. That is, if the number of newly infections per day decrease directly since the last date of the data, we then take the case as no second wave (the corresponding vaccination coverage is plotted as blue), otherwise, we say that there is another wave of the epidemic while the corresponding vaccination coverage is plotted as red.

It follows from Fig.6 and Fig.7 that the epidemics of UK, Indonesia, and Russia are still in the increasing phase while the daily reported case can peak in a short time when the AVC reaches around 100 percent in the two types of exit strategies or remaining the control interventions (no exit), i.e. the solid curves marked as black. That is, vaccination can shift the peak time forward. Furthermore, comparing the solid curves with different colours, we find that the vaccination can also significantly drop down the epidemic curves, particularly in terms of decreasing the peak value of the confirmed cases. For example, the peak number of the daily reported cases in Indonesia decreased from 2.1073 *∗* 10*^*6 to 5.1288 *∗* 10*^*3(more than 400 folds) when the AVC increases from 0 to 0.98. It follows Table 3 that the difference of the peak values with and without vaccination is relatively small in the strategy without exit compare to those with exit strategy of ES1 or ES2. This means that given strict NPIs, vaccination can decrease a relatively small number of the confirm cases.

As for the countries with the epidemics in the decreasing phase (China, Brazil, and US), the number of the daily reported cases may keep declining trend (no another wave) if they remain the control interventions (see the first row of Figs. 6-7) or have the alternative vaccination strategy with high coverage under the exit strategy of ES1 and ES2 (see the second and third row of Figs 6-7 or Table 3). However, if we rerelease the control interventions but without adding other interventions like vaccination, there may be a large second wave for these three countries. In more details, the daily reported cases can peak at 4.6992 *∗* 10*^*5 and 2.0917 *∗* 10^6^in Brazil under the exit strategy of ES1 and ES2 when there is no vaccination, respectively, as listed in Table 3. Particularly, although China has taken a very strict control interventions, there may be a large second wave with a peak value of 5.0987 *∗* 10*^*4 under the exit strategy of ES2 accompanied by the low coverage vaccination, i.e. the pink curve in the upper panel of Fig. 6. Similarly, from the panel of Fig. 7 for US, we can find that under both exit strategies of ES1 and ES2, when the vaccination coverage is low, then there will be always another wave of the epidemic while there will be no other wave when the AVC is high enough. This implies that there exists a critical coverage of vaccination which determines whether there is other wave of the epidemics.

Therefore, we further plotted the curves of the accumulative vaccination coverage 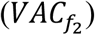 in Fig.8 as the vaccination rate increases in China, Brazil, and US. Taking the newly infected cases as an index, the red cycle means that with this vaccination rate and the AVC, there will be a second wave of the epidemics while the blue cycle means that there is no any other waves (i.e., the number of the newly infected cases decreases directly). Similarly, we find that there will be no second wave if the three countries can persistently remain the existing intensity of the control interventions. Once the contact rate returns back to the initial value (i.e. ES1), there will be another wave for both Brazil and US. To avoid the second wave, the minimum vaccination coverage should be around 0.92 and 0.19 for Brazil and US, respectively. In such case, there is still no second wave in China. If we further release the control interventions of quarantine (i.e. ES2), all these three countries will experience another wave given no vaccination. The minimum vaccination coverage in terms of avoiding another wave becomes around 0.41, 0.95, and 0.95 in China, Brazil, and US, respectively.

## Conclusion and discussion

The COVID-19 pandemic has posed a great threat to the global public health, and greatly influenced the normal-life of the residences all over the world. Although, various of non-pharmaceutical interventions (NPIs) helped to mitigate the epidemics, the urgent needs to reopen the economic development and resume the normal-life result in a more serious outbreak in many countries. Therefore, it’s critical to find an effective way to balance the controlling of the epidemics and the reopening of the economic development. Vaccine is expected to be an attractive and possible way to achieve this goal. In this study, utilizing two mathematical models (with and without COVID-19 vaccination), we showed how to design a dynamic vaccination regime to balance (replace) the dynamic exit of the NPIs and the controlling of the COVID-19 epidemics in terms of mitigating the epidemics, how much the vaccination coverage is needed to inhibit the possible waves.

Choosing six countries, including China, Brazil, Indonesia, Russia, UK, and US, as the examples, we firstly calibrated the model without vaccination by fitting the real data of daily reported cases and death cases. Further, considering the population resume the normal-life activity, the contact rate was assumed to return to its initial value in three stages, while the quarantine rate was assumed to either remain the estimated values (ES1) or drop to zero at the beginning of the vaccination (ES2). Re-defining the functions of quarantine rate *q*(*t*) and contact rate *c*(*t*) in the exit strategy of ES1 and ES2, and fixing the other parameters estimated from the model without vaccination (i.e. model (1)), we then fitted the model with COVID-19 vaccination (i.e. model (2)) to the same epidemic data. Consequently, we estimate the vaccination rates in the three stages and the accumulative vaccination coverages. The best fitting results provide the quantitative evidence that the designed dynamic vaccination program can be an effective method to balance the dynamic exit of NPIs in terms of mitigating the dynamics. Further, in terms of decreasing the cases, we showed that the early and fast implementation of the vaccination (at the increasing phase of the epidemics) will benefit the control of the epidemics a lot while vaccination initiating at the decreasing phase of the epidemic has a very limited influence. We also observed that the accumulative vaccination coverage needed to balance the exit of the existing NPIs in six countries are significantly depending on the strengthen of the control interventions. This quantitatively demonstrated the strengthen of the combined NPIs in the different countries are significant different.

In the second part, we assumed to vaccinate the population since the last data point of each country in three scenarios of exit strategies. We verified that vaccination can shift the peak time forward and also significantly drop down the peak value. Taking China, Brail, and US, as examples, we showed when the effective vaccination coverage is low, there can be another wave even with a larger peak. Note that, in the data section, we mentioned that we only included the epidemic data of US till September 30, 2020 while the epidemic lies on the decreasing phase of the second wave. In Fig. 9, we updated the data information in US till December 4, 2020 while the US are experiencing the third wave. It follows from Fig. 9 that our model can well describe and predict the development of the COVID-19 epidemics. Particularly, when we assume the contact rate return to the initial value while the quarantine rate just decreases to 0.23 (a weakened exit strategy of ES2), the prediction curve can fit the data very well, i.e. the curves marked as blue in Fig. 9. Note that, although the US tried to vaccinate the population, the coverage are still very low till December 04, 2020. This validation result well demonstrates our conclusion that there can be other waves if the NPIs exit but without an additional high level of vaccination coverage. On the other hand, we found that because the strengthen of the control interventions are significantly different among China, Brazil, and US, the AVC needed to inhibit a second wave is greatly different in the three countries. That is, a higher strengthen of control interventions is, a lower of the critical vaccination coverage should be. Furthermore, we showed that the combined NPIs can help to decrease the critical vaccination coverage to avoid a second wave. Therefore, it’s necessary to keep the normalized NPIs if the we have a limited access of vaccine.

**Fig. 9.**
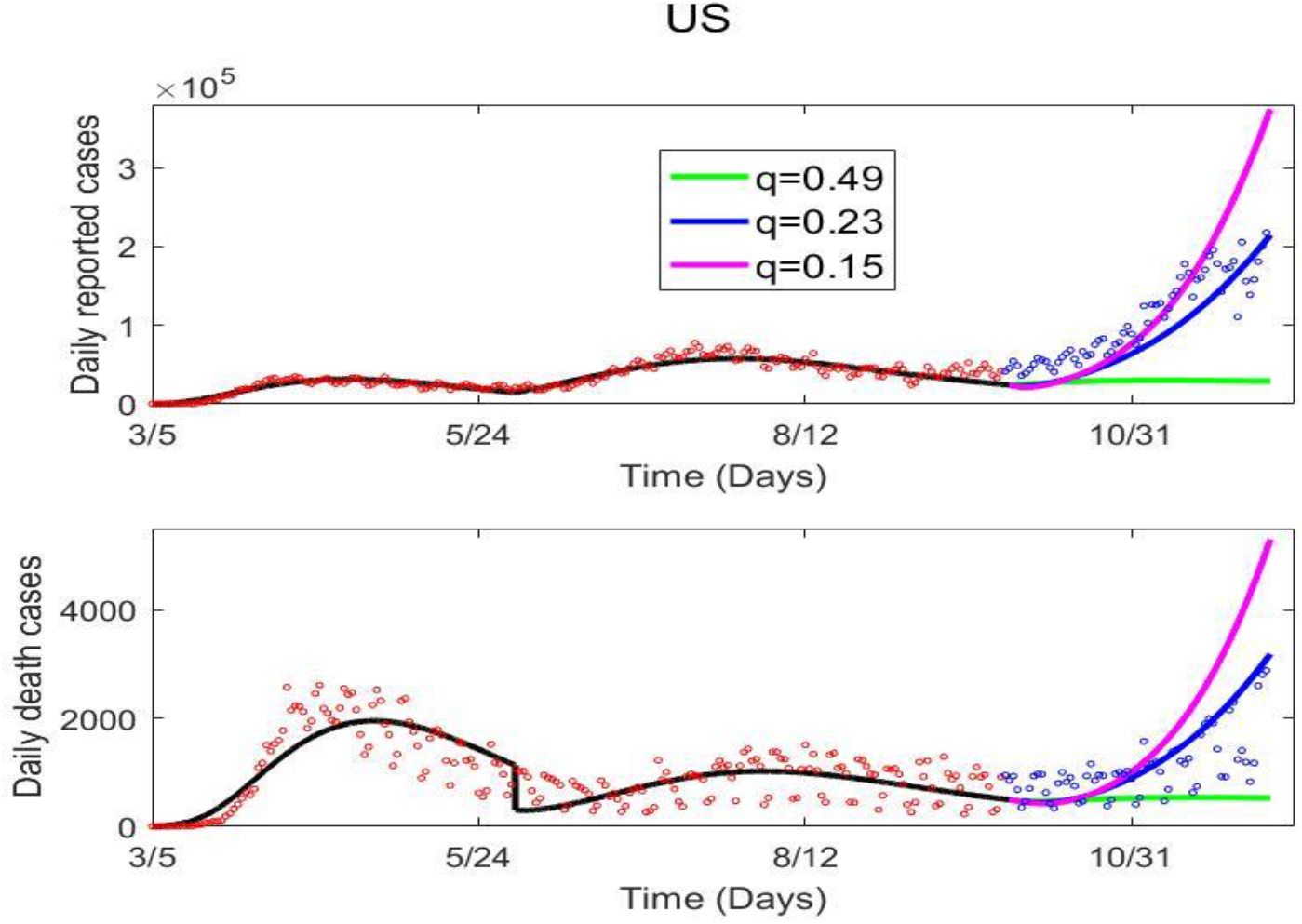
Predictions of the epidemics in US by assuming that the contact rate increasing to the initial value after Sep. 30, 2020 (the date of the last red cycle). Here the black curves are the best fitting curves in US. Note that, we only fitted the data of the red cycles while the data of the blue cycles are not fitted. The pink, blue and green curves are the predictions by choosing different quarantine rate after Sep. 30 with 0.49 is the estimated value for *q*_*b*_.

In conclusion, when we only have a limited access of the vaccine or the vaccine has a low efficacy but we want to resume the normal-life, it’s better to combine the normalized NPIs with the vaccinations aiming at inhibiting another wave of the COVID-19 epidemics. In other words, the exit of NPIs should be country or territory-based according to the strengthen of the NPIs, the accessibility, the affordability, and the efficacy of the vaccine.

## Data Availability

The epidemic data is public available.

## Author contributions

Conceptualization, BT, JW, YX, ST; validation and simulation, BT, PL, JY; data curation, BT, PL, JY; writing—original draft preparation, BT, YX, ST; writing—review and editing, BT, JW, YX, ST; All authors have read and agreed to the published version of the manuscript.

